# Predicting Protein Cascade Expression from H&E Images

**DOI:** 10.64898/2026.01.23.26344725

**Authors:** Alejandro Leyva, Abdul Rehman Akbar, M. Khalid Khan Niazi

**Affiliations:** Department of Pathology, The Ohio State University, 281 W Lane Ave, Columbus, OH 43210

**Keywords:** Artificial Intelligence, Omics, Genetics, Vision Transformers, Reverse Phase Protein Array

## Abstract

Protein expression within oncogenic or suppressive pathways is a hallmark indicator of oncogenesis. While traditional AI models in digital pathology attempt to predict singular proteins, there is a need to predict the downstream expression of proteins to indicate the propagation of signals. RNA expression provides novel information, but does not provide information about the downstream propagation of protein signals or whether those signals are functional. Using Reverse Phase Protein Array (RPPA) data with whole-slide images (WSIs) from the publicly available Cancer Genome Atlas Breast Adenocarcinoma dataset (TCGA-BRCA), we predict the expression of five key proteins identified from the apoptosis cascade, using DNA damage and repair (DDR) cascades as a biological control. Furthermore, we examine the performance of patch-level Vision Transformers (ViT) on the regression task, which was tested against the designed cellular-level ViT, CellRPPA. Our results demonstrate that patch-level vision transformers were unable to obtain statistically significant predictive results, achieving R-squared values ¡ 0.1 for all folds. In addition, CellViT obtained R-squared values ¿ 0.1 in all five test folds. We also show that morphologically indicative cascades, such as the apoptosis cascade, provide significantly higher performance compared to the DDR cascade.

## 1 Introduction

Breast adenocarcinoma is a highly documented disease and can be associated with a variety of genetic mutations that result in the propagation or suppression of protein signals [1]. Pathways can be intrinsically mediated, whereby the stimulus resulting in signal transduction is intracellular, or extrinsic. Within adenocarcinoma, common mutations in proteins such as TP53 result in the suppression of apoptotic signals by mutating the DNA-binding domain, resulting in dysfunctional proteins [2]. Other mutations, such as those in PIK3CA or MYC, result in the enrichment and propagation of the PIK3CT/AKT pathways by mutating kinase domains, leading to the overexpression of these proteins[3]. Apoptosis is responsible for regulating programmed cell death and is characterized by shrinkage, pyknosis, and reorganization of lipid structure [4]. The pathway is both intrinsically and extrinsically mediated by a combination of enzymes, receptors, and transcription factors[5]. Intrinsically mediated apoptosis occurs in two categorical fashions: negative, which is due to the absence of growth factors around a structure resulting in the triggering of cell death, or positive, which may be due to the presence of antigens, radiation, or hypoxia[6]. These changes result in the opening of inner mitochondrial pores due to lost membrane potential, preventing regular cell metabolism and causing the activation of BH3 proteins that detect metabolic stress[7]. As a result, BH3 proteins deactivate anti-apoptotic proteins such as BCL-2 and begin to activate apoptotic proteins such as BAK/BAX, resulting in the transduction of signals to XIAP (X-linked inhibitor of apoptosis protein), which then disinhibits the caspase family. The released caspases then catabolize the cell, resulting in cellular death[8].

The extrinsic pathway performs a similar function but is dependent on ligand–receptor binding of the TRAIL/FasL proteins, which results in the activation of caspases 8 and 10, which then activate the same caspases up to XIAP [9]. Common forms of extrinsic apoptosis include T-cell–mediated apoptosis and immune response [10].

The DNA damage and repair cascade is responsible for the repair of DNA in response to double-strand breaking or transcriptional errors. It has been demonstrated that lower expression of the DDR cascade is prognostically predictive within breast adenocarcinoma and has been predictive of chemotherapy sensitivity and response [11]. DDR applications within clinical oncology include testing for mutations in genes responsible for double-strand breaking repair, BRCA1 and BRCA2 (BReast CAncer Gene 1/2), and can be secondarily characterized by RNA expression. However, the choice of genes to use for prognostic value is still debated [12]. Moreover, the expression of DDR genes is not physically visible via an electron microscope but can indirectly result in abnormal cellular growth or polyploid cells [13]. Genes that are often used to characterize the DDR cascade include ATM, CHEK2, H2AFX, RAD51, and TP53, among others. The ataxia–telangiectasia mutated gene (ATM) regulates the response to double-strand breaking and senses DNA double-strand breaks [14]. RAD51 (radiation-sensitive protein 51) is responsible for the invasion of homologous DNA zones to allow for accurate and timely DNA repair and behaves as an ATPase [15]. CHEK2 (checkpoint kinase 2) is a protein responsible for producing a kinase that cleaves DNA regions and is a radiation-sensitive protein that prevents the cell from entering mitosis upon DNA damage [16]. CHEK2 has been shown to be a predictive biomarker for multiple organ cancers across Europe and North America [17]. TP53 is a transcription factor responsible for expressing the p53 antigen, as well as a protein that activates or inhibits apoptotic genes such as NOXA [18]. TP53 (tumor suppressor antigen 53) typically functions in multiple roles to suppress proliferative pathways and regulate cell cycling. H2AFX is responsible for regulating nucleosome formation and produces phosphorylative foci to recruit repair factors for double-strand breaks; the H2A family is also responsible for regulating chromatin accessibility and replication timing [19].

These cascades and their expression are typically predicted using RNA expression in the field of bioinformatics[20]. Within the field of digital pathology, gene expression models and molecular subtyping models have been developed using novel deep learning methods, whereby gene expression can be predicted from whole-slide image features [21]. More recently, multi-modal models integrating protein and genetic data are being developed, complementary to spatial transcriptomics and proteomics [22]. While RNA provides a perspective on cascade expression and prognosis, it does not indicate whether genes are translated and produced, or whether the proteins produced are functional [23]. Traditional models in the field have attempted to predict the expression of singular proteins or antigens that can be readily indicated from histology, including HER2 expression and EGFR [24, 25]; however, most models do not predict intracellular protein expression due to morphological ambiguity and poor generalization across cancers and external datasets. Multimodal models and comprehensive information on protein expression are required to understand protein functionalization for improved prognostic prediction. Information from proteomics contextualizes cellular behvavior and patterns in gene expression in systems biology, as cascades are highly interconnected and cross-talk mechanisms of action [26]. Misalignment between the extent of gene expression and the presence of proteins indicates translational or transcriptional dysfunction or deliberate inhibition.

Protein data are gathered from Reverse Phase Protein Array, which uses fluorescent antibodies that bind to the protein of interest [27]. The luminescence of the antibody indicates the expression of that protein, which is quantified by absorbance. The sensitivity and accuracy of RPPA depend on the affinity, specificity, and availability of proteins to bind to the provided antibodies and are conditionally accurate. Typically, multimodal analyses between RNA and RPPA within public datasets are not performed due to the time differential at which each assay is performed, rendering direct comparison between protein and RNA expression invalid. When using AI for expression prediction, information is extracted from region/patch level of whole slide images [28]. In recent years, higher resolution image embeddings have been developed to examine Images at the cellular level, and are untested for protein prediction [29]. Since Protein Cascades are generally visible at the cellular level, there is a need to investigate the ability of AI at both the patch level and the cell level to predict the expression of multiple proteins.

In this study, we predict the expression of multiple intracellular proteins in aggregate as a regression task from WSIs. We present a comprehensive algorithm, CellRPPA, inspired by CellEcoNet [30], to compete with conventional patch-level algorithms. This study offers two novel investigations: i) differences between cell-level and patch-level resolution in deep learning for protein expression prediction, and ii) the capacity for deep learning to predict morphologically ambiguous proteins, or proteins that are loosely indicated by histology. As digital pathology continues to expand, there is an ever-growing need for multimodal models and proteomics to provide a comprehensive perspective on disease progression.

## 2 Materials and Methods

From the publicly available Cancer Genome Atlas (TCGA) Breast Adenocarcinoma dataset (TCGA-BRCA), 919 RPPA samples were paired with whole-slide images in a 1:1 mapping, where cascades were defined using proteins. The apoptosis cascade was defined as the expression of BCL2, BAX, XIAP, and cleaved caspases 3 and 7. This was done to represent the activation of both intrinsic pathways demonstrated by BCL and BAX, as well as the expression of the encompassing caspases and regulatory inhibitors. The DDR cascade was used as a morphologically ambiguous protein cascade to compare against the performance of apoptosis, which is microscopically noticeable. The proteins chosen to represent the DDR cascade include H2AFX, CHEK2, TP53, TP53BP1, and ATM; however, this is acknowledged to be a limited representation of the DDR cascade and was chosen to represent hallmark genes associated with prognostic value in breast adenocarcinoma.

The RPPA scores for the relative intensity of each protein were then summed and z-scored across each protein. If any protein that was part of the cascade was not included within a sample, the sample was excluded entirely. The CellRPPA model is CellEcoNet reengineered with a regression head, and the GitHub repository for the model is included for both CellEcoNet and CellRPPA. Cell embeddings were extracted using Trident and CellViT++, at 20× magnification, and cell types were not determined.

The apoptosis and DDR tasks were performed separately, and training and evaluation took 150 hours for each task. The patch-level ViT uses a standard ViT-S/16 with cross-fold validation, where the DDR and apoptosis tasks were tested and validated separately and were not backpropagated jointly. Implementation details are available in the GitHub repository, and there are no specific modifications to the S/16 vision transformer. All parameterizations for both models are provided within anonymized shell scripts and should be able to run, provided that the necessary label files are available. Figure 1 shows the workflow for the study design, starting from the development of ground truth through the evaluation of each model. GO and KEGG analyses were performed using Bioconductor and ClusterProfiler in R 4.3.0, and all computational experiments were performed on the Ohio Supercomputer. All analyses were performed on the entire cohort, and the RPPA analytics were derived from the RPPA CSV using matplotlib, NumPy, and SciPy.

**Figure 1.**
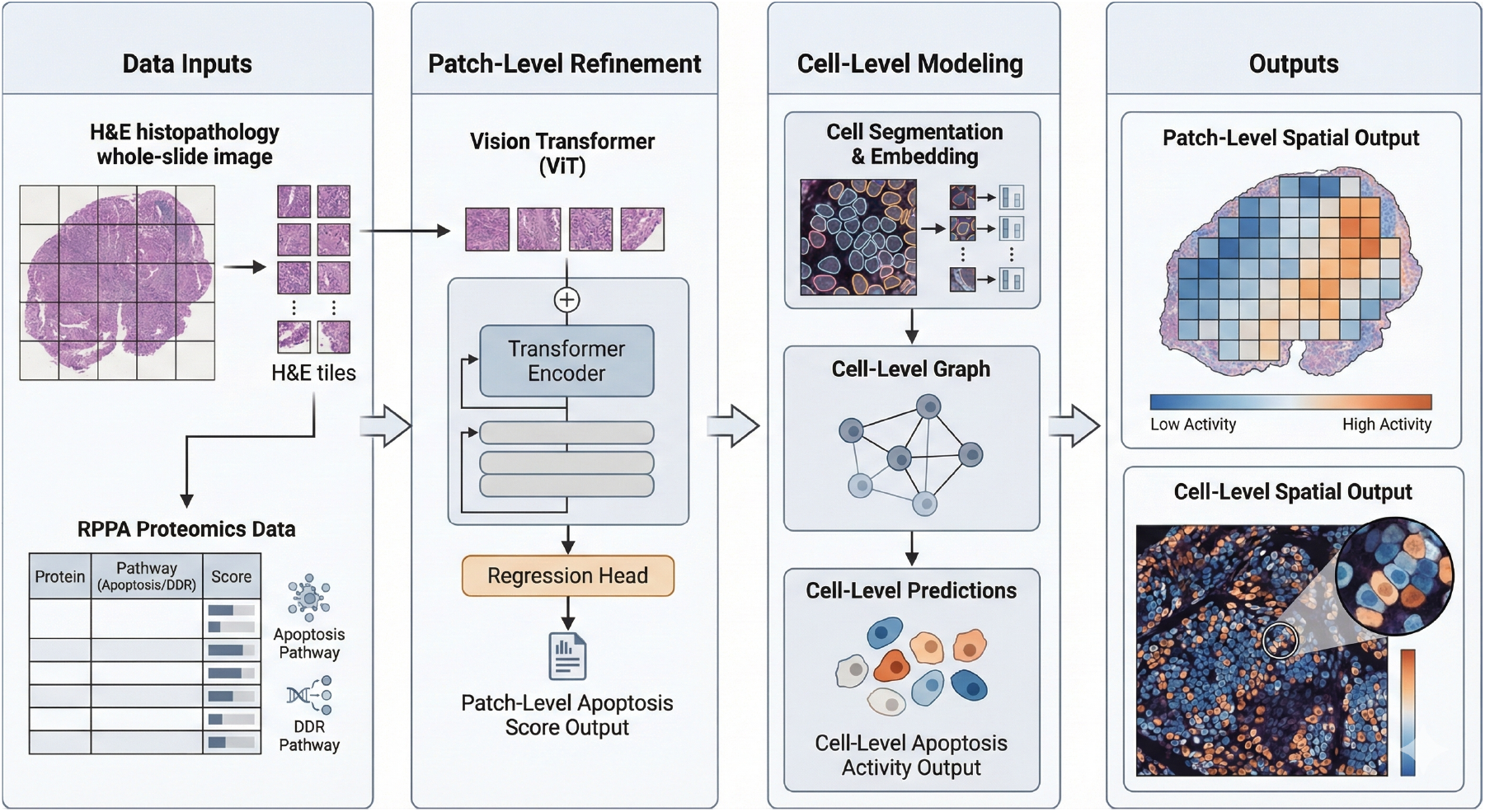
Overview of the cascade prediction framework. Whole-slide H&E images and RPPA-derived protein cascade scores serve as inputs. Patch-level modeling uses a Vision Transformer (ViT) to extract tile embeddings and regress pathway-level protein activity, while cell-level modeling performs cell segmentation, embedding, and graph construction to predict apoptosis activity at cellular resolution. The outputs illustrate spatially resolved pathway activity maps at both patch and cell scales, enabling comparison between coarse tissue-level predictions and fine-grained cellular cascade expression.

## 3 Results

The labels for the protein scores were assigned by case, and each case was assigned into fold-wise training, testing, and validation cohorts for evaluation over 100 epochs per fold. Initial analysis presents results on the expression of proteins defined within each sample as aggregated scores, as shown in Figure 2. Figure 2A shows the correlation heatmap of protein coexpression across each cascade. Proteins placed under the apoptosis cascade exhibit moderate coexpression among their counterpart proteins, demonstrating signal transduction within the cascade. BAX and XIAP show very low coexpression, as BAX must be inhibited in order for XIAP to be expressed. Interestingly, BCL2 and 53BP1 show stronger coexpression, above 50% Pearson’s correlation coefficient, while 53BP1 shows negative coexpression with other DDR genes within its defined cascade. 53BP1 also shows a strong correlation with XIAP expression, while exhibiting low coexpression with BAX. BCL2 and BAX show low protein expression correlation, which can be inferred from the biological roles of these proteins.

**Figure 2.**
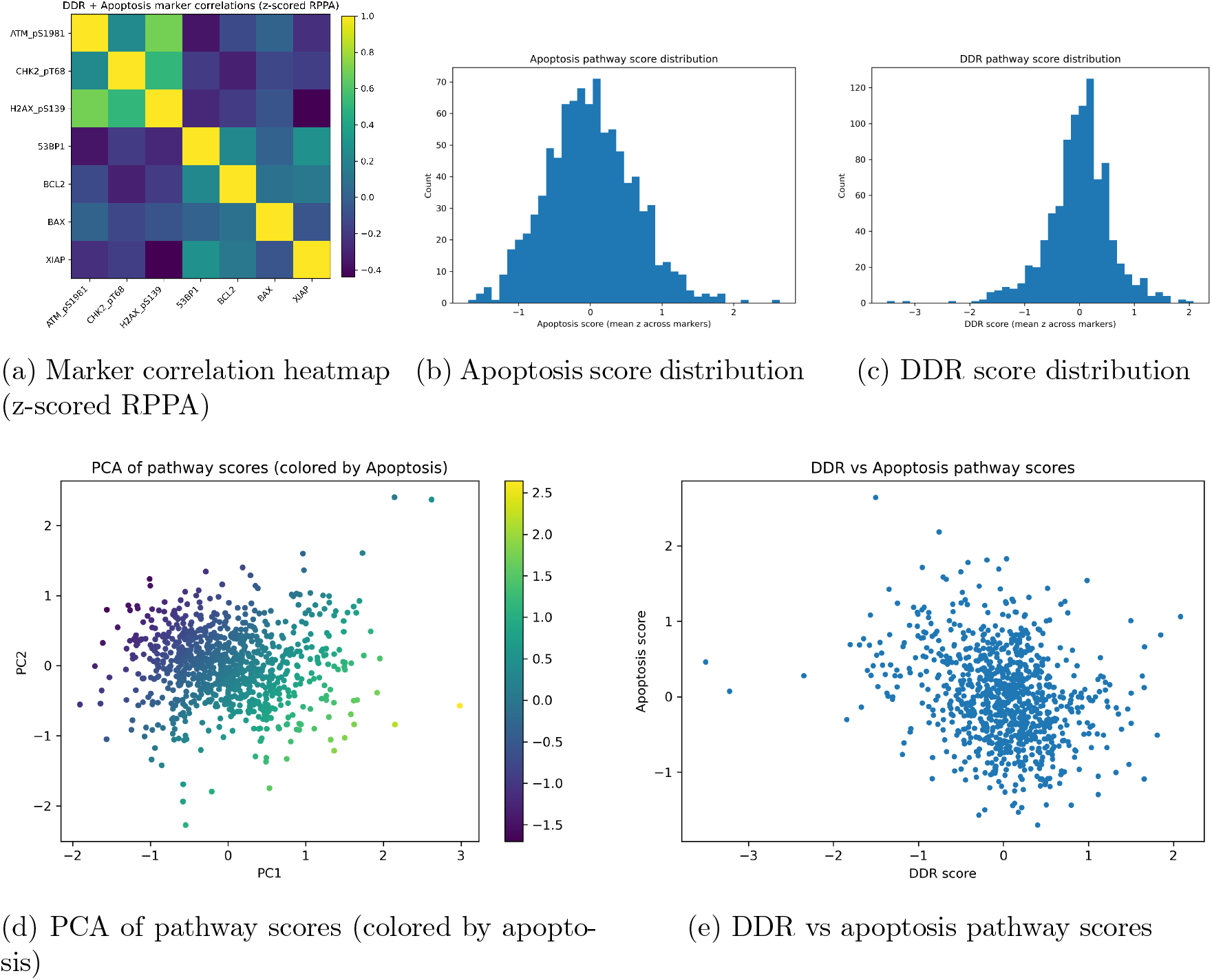
Proteomics and label-level analytics for apoptosis and DDR RPPA-derived pathway scores. Top row: inter-marker correlation structure and pathway score distributions. Bottom row: low-dimensional structure of pathway scores and cross-cascade correlation (control relationship between DDR and apoptosis).

Within the DDR cascade, 53BP1 shows lower coexpression with DDR-defined genes, including ATM, CHK2, and H2AX. The coexpression correlation between ATM and all other DDR genes, except for 53BP1, is remarkably strong, exceeding 80% with H2AX and 50% with CHK2. The correlation heatmap shows little overlap between the cascade expressions of DDR and apoptosis and often demonstrates negative coexpression between cascades. There is a small positive correlation between BAX and ATM, while most other correlations are below zero.

Figure 2B shows the standard Gaussian distribution of apoptosis pathway scores, with no observable skew and larger variance from the median. Figure 2C shows the DDR score distribution, which maintains a standard Gaussian distribution with outliers that are three standard deviations away from the mean, attributed to the absence of expression of any classified DDR gene. Figure 2D shows the PCA of pathway scores using UMAP projections for each sample, where samples that are closer together have similar protein expression profiles. A higher frequency of samples cluster toward minimal variance, while outliers, or cases with higher apoptosis cascade scores, are more dispersed and exhibit greater variance in protein expression, or markedly higher expression of proteins that are typically less expressed.

To visualize the relationship between DDR scores and apoptosis, Figure 2E shows a plot that classifies samples by their respective scores. In general, samples with higher apoptosis scores tend to have lower DDR scores, and vice versa. However, a large proportion of samples with apoptosis scores near zero also have DDR scores near zero, suggesting an inverse relationship.

The results for the patch-level ViT on the morphologically ambiguous DDR cascade, used as a control, are shown in Table 1. The model failed to obtain statistically significant correlations in three folds using False Discovery Rate (FDR) correction on the Spearman correlation for non-linear correlational analysis. Fold 2 completely failed to obtain any Pearson or Spearman correlation as a result of convergence, while the MAE remained relatively high. MSE did not consistently decline across folds, and the MAE remained stagnant across all folds. The patch-level ViT is shown to predict close to no variance in protein expression within the DDR cohort across all five folds. In addition, the Spearman and Pearson values were nearly identical in some folds, indicating similarity in morphological correlation.

**Table 1:** Performance metrics for DDR pathway prediction across folds. Missing values are reported as zero.

| Fold | Pearson $r$ | Pearson $p$ | Spearman $\rho$ | Spearman $p$ | MAE | MSE | $R^2$ | FDR sig |
| --- | --- | --- | --- | --- | --- | --- | --- | --- |
| 1 | 0.1479 | 0.0870 | 0.1513 | 0.0799 | 0.4219 | 0.3009 | 0.0135 | False |
| 2 | 0 | 1.0000 | 0 | 1.0000 | 0.4437 | 0.4088 | -0.0011 | False |
| 3 | 0.1243 | 0.1510 | 0.1803 | 0.0364 | 0.4286 | 0.3674 | -0.0199 | False |
| 4 | 0.2507 | 0.0032 | 0.2507 | 0.0032 | 0.4106 | 0.3170 | -0.0019 | True |
| 5 | 0.2169 | 0.0115 | 0.1418 | 0.1010 | 0.4156 | 0.3497 | 0.0320 | True |

The results for the patch-level ViT’s prediction of apoptosis cascade expression are shown in Table 2. The results are noticeably worse for apoptosis, with only one of the five folds showing significant correlations and three of the five folds showing no correlation at all. The MAE also increased relative to the DDR cascade prediction, exceeding 0.5 in some folds, while the typical range of values is between 0 and 1. The MSE did not remain steady and did not consistently improve across folds; Fold 1 demonstrated the best MSE, which then steadily worsened. The model failed to explain any variance in protein expression and, in Fold 3, produced slightly negative values, demonstrating inadequate capability to predict apoptosis cascade expression, despite apoptosis being morphologically visible and the known correlation between protein expression and cascade expression.

**Table 2:** Performance metrics for apoptosis pathway prediction across folds. Missing values are reported as zero.

| Fold | Pearson $r$ | Pearson $p$ | Spearman $\rho$ | Spearman $p$ | MAE | MSE | $R^2$ | FDR sig |
| --- | --- | --- | --- | --- | --- | --- | --- | --- |
| 1 | 0.0392 | 0.6515 | 0.0445 | 0.6082 | 0.4184 | 0.2955 | 0.0003 | False |
| 2 | 0 | 1.0000 | 0 | 1.0000 | 0.5120 | 0.4067 | -0.0116 | False |
| 3 | 0 | 1.0000 | 0 | 1.0000 | 0.4836 | 0.3702 | -0.0330 | False |
| 4 | 0.2236 | 0.0089 | 0.2530 | 0.0030 | 0.5302 | 0.4424 | 0.0029 | True |
| 5 | 0 | 1.0000 | 0 | 1.0000 | 0.4610 | 0.3313 | -0.0018 | False |

**Table 3:** Patch-level apoptosis prediction performance across five-fold cross-validation. Reported metrics include Pearson correlation, Spearman correlation, mean absolute error (MAE), mean squared error (MSE), and coefficient of determination (*R*^2^).

| **Split** | **Fold** | **Pearson** | **Spearman** | **MAE** | **MSE** | **R<sup>2</sup>** |
| --- | --- | --- | --- | --- | --- | --- |
| 5*Train | Fold 1 | 0.5734 | 0.5626 | 0.3811 | 0.2321 | 0.3257 |
|  | Fold 2 | 0.6147 | 0.6097 | 0.3846 | 0.3421 | 0.3773 |
|  | Fold 3 | 0.6009 | 0.5733 | 0.3808 | 0.2315 | 0.3572 |
|  | Fold 4 | 0.6072 | 0.5859 | 0.3906 | 0.2327 | 0.3926 |
|  | Fold 5 | 0.5969 | 0.5819 | 0.3737 | 0.2332 | 0.3515 |
| 5*Validation | Fold 1 | 0.5000 | 0.4508 | 0.4771 | 0.3584 | 0.2446 |
|  | Fold 2 | 0.4300 | 0.4274 | 0.4144 | 0.2821 | 0.1708 |
|  | Fold 3 | 0.4564 | 0.4579 | 0.4806 | 0.3551 | 0.2064 |
|  | Fold 4 | 0.4736 | 0.4669 | 0.4419 | 0.3083 | 0.1601 |
|  | Fold 5 | 0.5564 | 0.5754 | 0.4326 | 0.2915 | 0.3068 |
| 5*Test | Fold 1 | 0.4982 | 0.5199 | 0.4177 | 0.2800 | 0.2414 |
|  | Fold 2 | 0.3770 | 0.3574 | 0.4645 | 0.2332 | 0.1222 |
|  | Fold 3 | 0.4629 | 0.4688 | 0.4227 | 0.2855 | 0.1659 |
|  | Fold 4 | 0.4454 | 0.4439 | 0.4469 | 0.3135 | 0.1648 |
|  | Fold 5 | 0.5084 | 0.4755 | 0.4220 | 0.2783 | 0.2484 |

Gene ontology analysis, cellular component analysis, and gene functional analysis were performed to understand the morphological characteristics that the model would learn from histological patterns, providing in-depth analysis of which histological features correspond with gene expression, and thus protein expression, and offering insight into the statistical patterns the model would learn. As shown in Figure 3, an initial gene ontology analysis was performed on the proteins observed in the apoptosis cascade. Figure 3A demonstrates that all genes chosen from the apoptosis cascade correspond directly with immune homeostasis associated with conventional external pathways, as well as regulation of cellular population. Gene ratio corresponds to the percentage of genes that possess a given function, and there is functional overlap across the apoptosis cascade among three of the apoptosis-related proteins, specifically in response to UV light, homeostasis, leukocyte apoptosis, and related processes.

**Figure 3.**
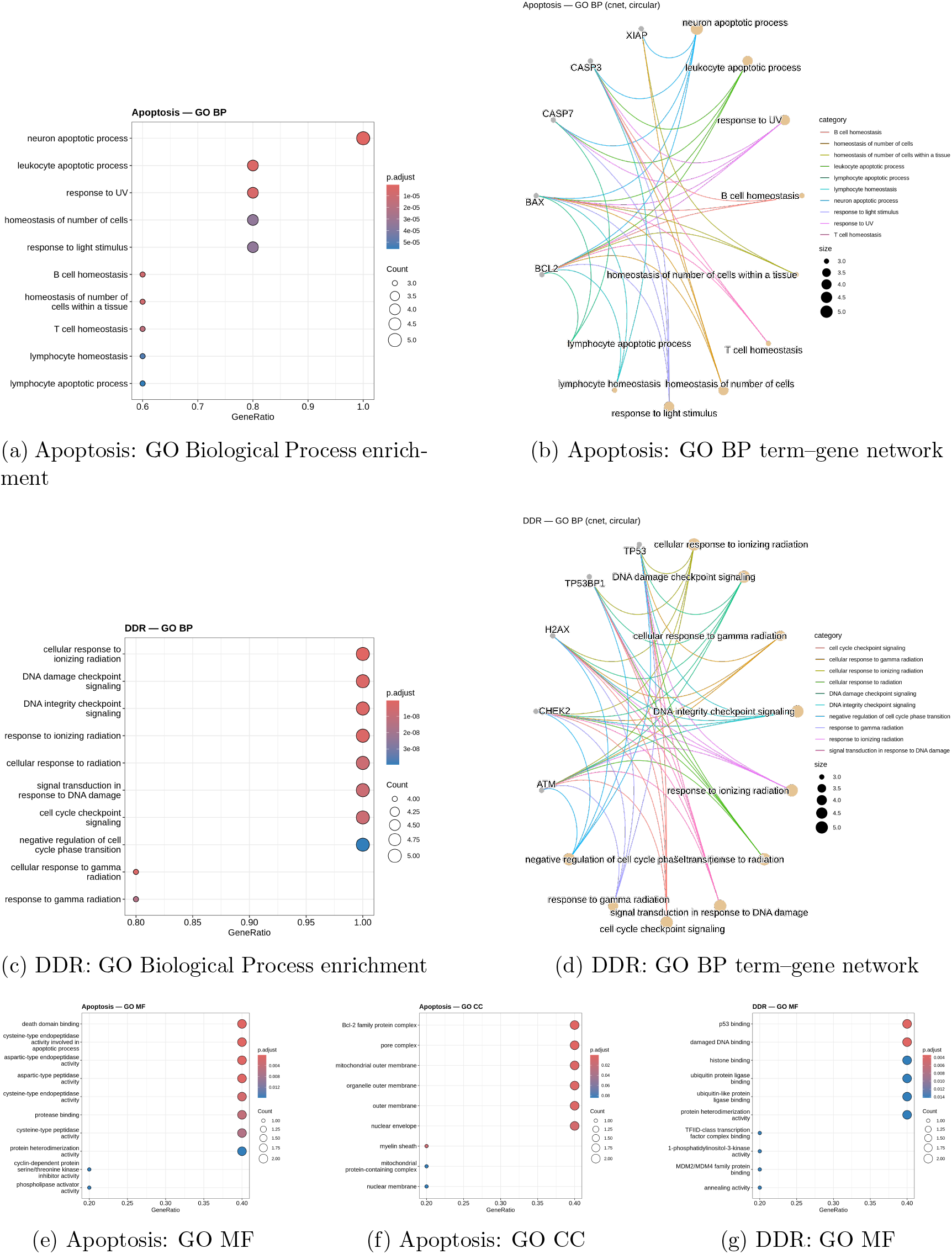
Gene Ontology (GO) functional context of the apoptosis and DNA damage response (DDR) protein sets. GO Biological Process (BP) enrichment demonstrates functional coherence of each cascade, while circular term–gene networks illustrate the relationships between enriched processes and contributing proteins. GO Molecular Function (MF) and Cellular Component (CC) analyses are shown for contextual support.

It is important to note these functions, as protein expression was extracted from bulk RPPA rather than single-cell data; therefore, there is no feasible way to associate protein expression with individual cell types. Neuronal apoptosis is a shared function across all five proteins and may be relevant to peripheral neuron degeneration within breast adenocarcinoma. The mappings of these apoptosis genes and their functions are shown in Figure 3B, where most genes exhibit overlapping functions, demonstrating that they exist within the same functional and cascade domains and further justifying the aggregation of proteins, as both functional overlap and coexpression are observed. Response to ultraviolet and visible light ontologically corresponds to BAX, CASP7, CASP3, and BCL2. T-cell homeostasis corresponds to BAX, BCL2, and caspase 3 only. Lymphocyte apoptosis corresponds to BAX, BCL2, and caspase 7, while lymphocyte homeostasis corresponds to BAX, BCL2, and CASP3, but not CASP7. Leukocyte apoptotic processing contains a duplicate entry in the GO analysis, resulting in redundancy. Regardless, these apoptotic proteins demonstrate high statistical significance for response to stimuli, homeostasis, and leukocyte apoptosis.

Gene ontology analysis performed for the five proteins selected for the DDR cascade is shown, where there is high statistical significance across all genes for genome integrity and response to radiation. There is overlap between the functions of apoptosis and DDR, as both respond to forms of radiation outside the visible spectrum. Since there is overlap among four of the genes used in the DDR cascade, it can be inferred that all proteins selected for prediction within the DDR cascade are viable and coexpressed. Because these proteins share DNA-preserving functions, it is also clear that they are not directly indicated in histology, but may result in various phenotypes depending on the upregulation or downregulation of DDR. The functional map shown in Figure 3D demonstrates significant overlap among the functions described in the GO analysis, as expected; however, only four of the selected genes correspond to response to gamma radiation, one of which is cellular and another related to signaling. TP53BP1 shows no role in response to gamma radiation, while all other functions overlap.

Molecular function analysis for the apoptosis cascade, shown in Figure 3E, demonstrates that the genes and proteins used for cascade prediction span a range of functions encompassing apoptosis. While peptidase activity for cysteine and other amino acids is represented by caspases 3 and 7, other functions, including death domain binding and protease binding, are regulated by BAX, BCL2, and XIAP, respectively. There is lower statistical significance for protein heterodimerization, which is a function of TP53BP1 and p53 and is required for activation of apoptosis-related proteins. Cyclin and phospholipase activator activities correspond to H2AX. Figure 3F presents the cellular component analysis of each protein and confirms that all proteins participate in both intrinsic and extrinsic pathways related to replicative and metabolic stress and TRAIL/FASL binding. Finally, Figure 3G summarizes the molecular functions of the five DDR proteins, which encompass ubiquitin binding and DNA repair, covering both intrinsic and extrinsic pathways. Overall, these results demonstrate that the predicted proteins encompass the apoptosis and DDR pathways, have meaningful functional indicators, and cover key components of pathway mechanisms for both cascades.

The results for CellRPPA’s prediction of the apoptosis cascade are shown in Table 4, demonstrating drastic improvement over the patch-level ViT, which indicates that higher resolution can be advantageous for the task. For each fold, the epoch with the lowest validation loss was chosen to represent performance across folds. All folds explained at least 10% of the variance in the protein prediction task, while a substantial portion of folds in both the test and validation sets exceeded 20 or 30%. While the MAE remains consistent across cohorts, there is controlled variance across folds. The Pearson and Spearman correlation coefficients across all cohorts were above 40%, with the exception of one fold in the test set. The MSE remained consistent relative to MAE values across cohorts and folds, ranging between 29–35, with the exception of two instances in the test set.

**Table 4:**
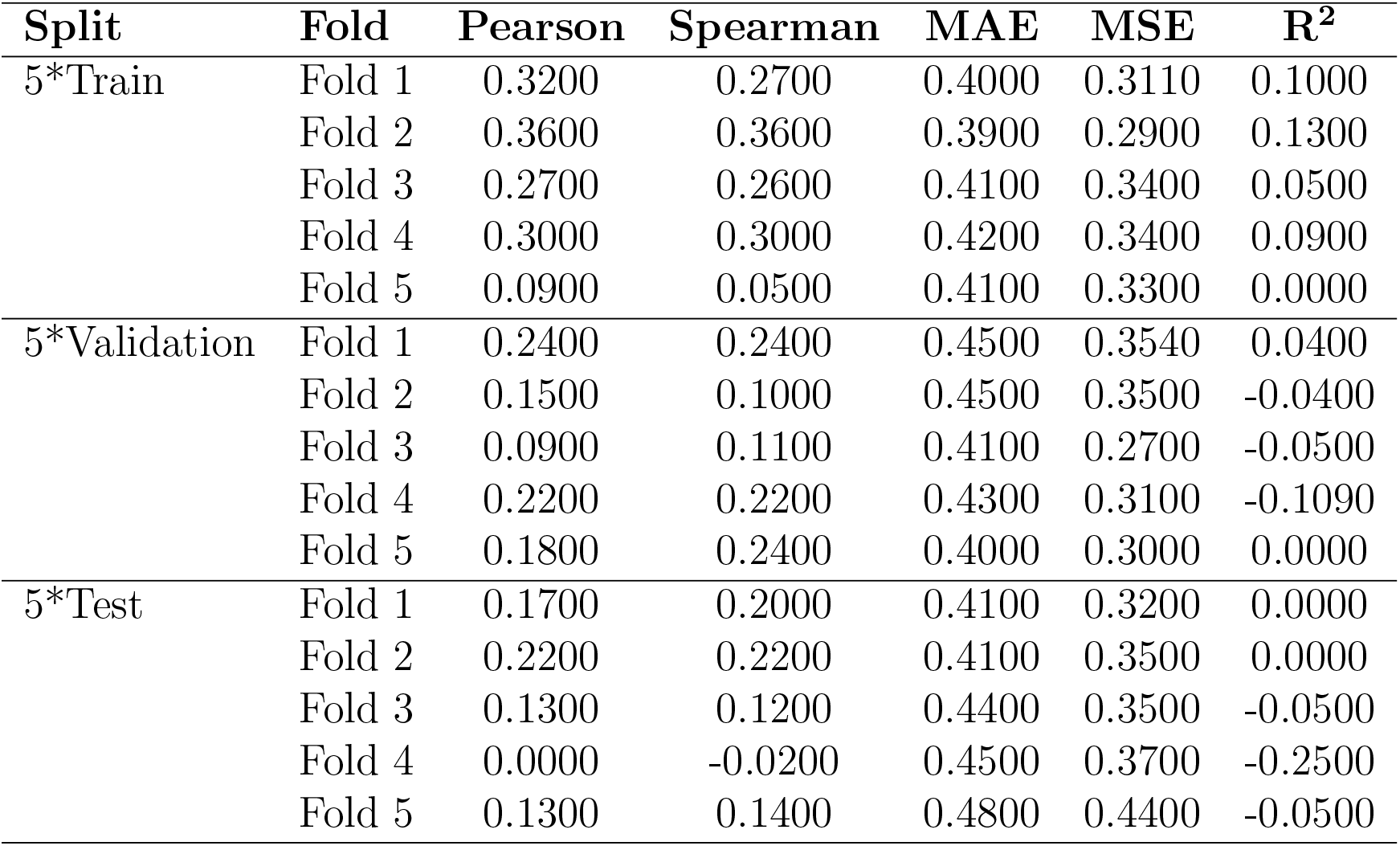
Patch-level DNA damage response (DDR) prediction performance across five-fold cross-validation. Metrics include Pearson correlation, Spearman correlation, mean absolute error (MAE), mean squared error (MSE), and coefficient of determination (*R*^2^).

| **Split** | **Fold** | **Pearson** | **Spearman** | **MAE** | **MSE** | **$R^2$** |
| --- | --- | --- | --- | --- | --- | --- |
| 5*Train | Fold 1 | 0.3200 | 0.2700 | 0.4000 | 0.3110 | 0.1000 |
|  | Fold 2 | 0.3600 | 0.3600 | 0.3900 | 0.2900 | 0.1300 |
|  | Fold 3 | 0.2700 | 0.2600 | 0.4100 | 0.3400 | 0.0500 |
|  | Fold 4 | 0.3000 | 0.3000 | 0.4200 | 0.3400 | 0.0900 |
|  | Fold 5 | 0.0900 | 0.0500 | 0.4100 | 0.3300 | 0.0000 |
| 5*Validation | Fold 1 | 0.2400 | 0.2400 | 0.4500 | 0.3540 | 0.0400 |
|  | Fold 2 | 0.1500 | 0.1000 | 0.4500 | 0.3500 | -0.0400 |
|  | Fold 3 | 0.0900 | 0.1100 | 0.4100 | 0.2700 | -0.0500 |
|  | Fold 4 | 0.2200 | 0.2200 | 0.4300 | 0.3100 | -0.1090 |
|  | Fold 5 | 0.1800 | 0.2400 | 0.4000 | 0.3000 | 0.0000 |
| 5*Test | Fold 1 | 0.1700 | 0.2000 | 0.4100 | 0.3200 | 0.0000 |
|  | Fold 2 | 0.2200 | 0.2200 | 0.4100 | 0.3500 | 0.0000 |
|  | Fold 3 | 0.1300 | 0.1200 | 0.4400 | 0.3500 | -0.0500 |
|  | Fold 4 | 0.0000 | -0.0200 | 0.4500 | 0.3700 | -0.2500 |
|  | Fold 5 | 0.1300 | 0.1400 | 0.4800 | 0.4400 | -0.0500 |

Table 5 shows the prediction performance for the DDR cascade using CellRPPA, which demonstrated worse values in comparison to apoptosis prediction performance. Relative to the patch-level tasks, CellRPPA showed no improvement over the patch-level ViT in terms of explained variance. Similar to the patch-level ViT, the Spearman and Pearson correlation coefficients fall within the same range, and the PCC values are similar, if not equal, to the Spearman coefficients. The MAE remained within a consistent range, as observed in the other predictive tasks, and was relatively stable across folds but did not exhibit a noticeable decrease.

The results demonstrate the failure of patch-level ViTs on multi-protein prediction for apoptosis and DDR. In contrast, for cell-level ViTs (CellRPPA), the model succeeds in predicting apoptosis but fails on DDR, reflecting the histological ambiguity of these proteins. Validation of protein choices for both DDR and apoptosis demonstrates that the selected proteins behaved within the same cascade and function and also exhibited noticeable coexpression. It is also noted that samples with lower DDR expression had higher apoptosis protein expression as measured by RPPA.

## 4 Discussion

Analysis of breast adenocarcinoma suggests that, in general, there is an inverse relationship between DDR expression and apoptosis expression, which is validated by the correlation heatmap shown in Figure 2A, indicating that a functional basis exists within breast adenocarcinoma. Previous studies indicate that the apoptosis and DNA damage response cascades are tightly coupled rather than independent. DDR signaling functions upstream to assess genomic damage and, when repair fails, promotes apoptotic commitment. Following irreversible activation of executioner caspases, key DDR components are cleaved, effectively terminating DNA repair processes [31]. While a few outliers exist in each cohort, the analysis suggests that most proteins exhibit similar levels of expression within the cohort.

Gene ontology analysis performed on the apoptosis and DDR genes suggests that the selected genes are aligned and that each protein exhibits corresponding and overlapping functions. Apoptotic signaling has been extensively characterized in response to UV-induced cellular stress, whereas the DNA damage response has been classically studied in the context of ionizing and gamma radiation–induced double-strand breaks. However, both pathways are activated across radiation modalities, with relative dominance depending on damage type, dose, and cellular context [32]. This relationship is supported by the correlation heatmap shown in Figure 2A. These results demonstrate the effectiveness of cascade-based scoring and support the viability of DDR as a morphological control against apoptosis, forcing the model to learn different histological patterns during training. The overlapping functions suggest that these processes are contextually distinct: apoptosis governs the termination of cellular function following sufficient cellular or metabolic damage, whereas the DDR cascade is responsible for initiating repair and maintaining thresholds for DNA repair.

Apoptosis is well known to be visible under compound or fluorescent microscopy, and it is notable that the patch-level ViT fails to detect and predict cascade expression and performs markedly worse than on the DDR cascade. Performance on the DDR cascade in both trials demonstrates that prediction is not suitable at either resolution, likely due to the inherent morphological ambiguity of DDR expression in histology. DDR expression may reflect replicative stress, aneuploidy, and other phenomena that can manifest in multiple visible forms across different cell types. In contrast, apoptosis has been observed to exhibit a set of well-characterized behaviors that are similar across most cell types. It is notable that the patch-level ViT does not provide sufficient resolution to statistically learn the patterns that characterize apoptosis. These results suggest that cascade prediction is more successful for proteins with histologically visible behaviors and that higher-resolution representations can be advantageous for such cascades.

## 5 Limitations

While this study uses the term “cascades” to describe multi-protein prediction, each cascade represents a characterized set of hallmark proteins from canonically observed pathways. Full representation of each cascade would require a substantially larger protein cohort, which would introduce additional noise and increase prediction difficulty. We also note that the model choices used for the patch-level ViT are limited and not fully generalizable, and similar limitations apply to CellViTs. Future studies should derive more generalizable conclusions across omics prediction by conducting broader evaluations of currently available models to assess the advantages of cell-level ViTs on omics tasks. In this study, minimal architectures were used to establish baseline performance for multiprotein prediction.

## 6 Conclusion

Using cell-level ViTs and patch-level ViTs, we predict the expression of hallmark proteins within cascades and assess the advantages and disadvantages of each modeling approach.

Our results show that cascade prediction is a viable task and can provide insight into molecular, genetic, and cellular functions and responses. Overall, we consider the performance of CellRPPA successful in comparison to other omics-prediction tasks, though improvement in MAE is needed for viability. We demonstrate that patch-level ViTs do not outperform CellRPPA in microscopically visible cascades and that CellRPPA does not provide an advantage over patch-level models for morphologically ambiguous proteins within the DDR cascade. It can be concluded that CellRPPA does not provide an advantage over patch-level ViTs for cascade prediction when targeting morphologically ambiguous proteins. Future studies should extend this analysis to additional cascades to better understand the integration of deep learning–derived proteomics into clinical informatics. In addition, cross-cohort analyses are needed to demonstrate generalization across cancer types.

## 7 Ethics Statement

All datasets are public and anonymized.

## 8 Author Contributions

All work was performed by the first author, CellEcoNet and embeddings were developed by the second author, with supervision by the third author.

## 9 Conflict of Interest

No conflicts of interest have been declared. The authors have reviewed the manuscript and have approved submission.

## 10 Funding

The project described was supported in part by R01 CA276301 (PIs: Niazi and Chen) from the National Cancer Institute, Pelatonia under IRP CC13702 (PIs: Niazi, Vilgelm, and Roy), The Ohio State University Department of Pathology and Comprehensive Cancer Center. The content is solely the responsibility of the authors and does not necessarily represent the official views of the National Cancer Institute or National Institutes of Health or The Ohio State University.

## 11 Data Availability

datasets and diagnostic slides are publically available and anonymized. CellEcoNet gihub is available here: https://github.com/AI4Path-Lab/PathRosetta. CellRPPA and preprocessing code is available here: https://github.com/Alejandro21236/CellRPPA.

## Notes

### Competing Interest Statement

The authors have declared no competing interest.

